# Analysis of Repeated Measurements of HIV Viral Load as a Continuous Variable while Accounting for Missing Values

**DOI:** 10.1101/2024.01.26.24301856

**Authors:** Susan Habert Sendege, Saint Kizito Omala, Symon Peter Wandiembe, Isaac Dumba

## Abstract

Recent HIV research predominantly uses Single Measure Frameworks (SMF), focusing solely on the latest viral load data and overlooking missing values. This study explored repeated measures frameworks to assess factors affecting viral load copies while accounting for missing data. The analysis involved 1670 records of HIV patients,using the generalized linear mixed models (GLMM). All variables, except for treatment regimen changes and adherence rating, were recorded at patients’ treatment enrollment. A GLMM was applied to data before and after imputation accounting for the repeated nature of the HIV viral load copies over time. The best-fitting model, selected for discussion, was the GLMM fitted to multiply imputed data. Gender and adherence rating did not significantly affect viral load copies. The analysis included other variables such as patient age, marital status, treatment duration, WHO clinical stages, and facility ownership. Results show that viral load copies were higher among currently or formerly married individuals (*β* = 0.49, 0.30; *SE* = 0.042, 0.052; *p* = 0.0000). In contrast, viral load copies were lower for patients with longer treatment durations (*β* = −0.01; *SE* = 0.001; *p* = 0.0000) and those receiving treatment at a private facility (*β* = −0.196; *SE* = 0.077; *p* = 0.0000). The study highlights the significance of recognizing repeated data patterns in longitudinal settings and addressing missing values in health research. It proposes a similar investigation in controlled environments to evaluate SMF and RMF in presence of missing values.

**Author summary:** This work was conceptualized by, Susan Habert Sendege. The author was responsible for the initial drafting of the paper, including the analysis, discussion, conclusions, and recommendations. Saint Kizito and Symon Peter played a pivotal role in shaping the study and all provided numerous hours of time for proofreading and editing.

## Introduction

Over four decades since the outbreak of HIV, the virus continues to be a danger to humanity [10]. According to the World Health Organisation (WHO), there were 38.4 million HIV infections worldwide in 2021, with 1.5 million new cases [16]. Sub-Saharan Africa (SSA) remained the region most affected by the virus, with a range of 470 000–930 000 new cases occurring in SSA [15]. Thanks to the development of anti-retroviral therapy (ART), many infected individuals can now live longer and with lower transmission risks than in the early days of the virus [6]. In HIV treatment and research, a direct measure of viral replication is viral load (VL), which is quantified as the number of copies of HIV ribonucleic acid (RNA) per milliliter of blood [5]. Regular viral load assessments are conducted in HIV patients with a recommended window of six months after starting Antiretroviral Therapy (ART). The goal is to assess treatment response and identify necessary modifications [17].

Statistical analysis is commonly used in health programs to draw conclusions from repeated measures data. In such a case, historical techniques using single observations may be inappropriate, as they neglect the intrinsic link between measurements over time [7]. Broadly, there are two frameworks for analyzing repeated measures data: that is, Single Measures Framework (SMF) and Repeated Measures Framework (RMF) [13]. Single measures with continuous or numeric observation and single measures with binary observation are two approaches to modeling under SMF. On the other hand, RMF treats all observations as obtained from the patients in the continuous case, while binary RMF models group information based on measurements at a predetermined cutoff [13]. SMF has been criticized for information loss, underestimation of variation, and suppressing linear relationships [2, 13]. As such, when using SMF, especially with binary categorization, the outcome variable may not be efficiently examined, and the decisions made may not be optimally supported.

When dealing with health data, missing data is also inevitable and should be handled with care for reliable results [9, 12]. However, numerous investigations on the quality of life of HIV patients have largely adopted the complete case analysis, which has the same effect as SMF. This study evaluated the candidature of selected factors under SMF and RMF, multiple imputation (MI), and complete case analysis while handling viral load copies as continuous variables. The study hypothesed that models fitted on multiply imputted data performs better than those fitted under the assumption of complete case analysis. Of interests to the study was patient gender and adherence scores where the two factors were assummed to be key explainers of viral load copies.

The rest of the paper is arranged as follows, Section two presents the methodology adopted in the study detailing the data sources, data management, and description of the variables included in the analysis. Section three presents the rsults at two levels (descriptive and inferential level) and the distribution of the random effects term. The section ends with conclusions and recommendations arising from the analysis.

## Materials and methods

### Study Design

The study was a retrospective longitudinal design in which the repeated measurements of viral load taken from a patient since initiation on ART were used as the dependent variable. The study was based on HIV data for a period of six years, from January 2016 to December 2021. The period of study was based on Uganda’s adoption of the test-and-treat policy in the year 2016.

### Data Source and Description

The study was based on secondary data obtained from health facilities providing HIV treatment and care in Mukono Municipality, following the formal approval of the District Health Officer (DHO). For some health facilities, data was obtained up to the tenth visit, and for others, data was only available up to the sixth visit. To strike a balance in the number of copies, all visits above the sixth were dropped from the study.

### Target Population and Sample Size

The target population was all HIV patients who had been initiated on ART between January 2016 and December 2021. The period was chosen based on the test-and-treat policy, which was adopted by Uganda’s Ministry of Health (MoH) in the year 2016 [11]. Data on 6040 individual patients were obtained and exposed to the inclusion and exclusion criteria explained below.

#### Inclusion Criteria

All patients who had their viral load measured at least twice were included in the analysis. Secondly, all patients with no detectable viral load copies at baseline (first visit) were excluded. To control the number of outliers in the analysis, all individuals with viral load copies greater than three standard deviations of the mean at a given visit *t hat is*, (*y*_*i j*_ ≥ 3σ_*y j*_) were also excluded from the study. The resulting sample was 1670, and these were the observations eligible for further analysis. **Figure 1** shows step-by-step inclusion and exclusion criteria.

**Fig 1.**
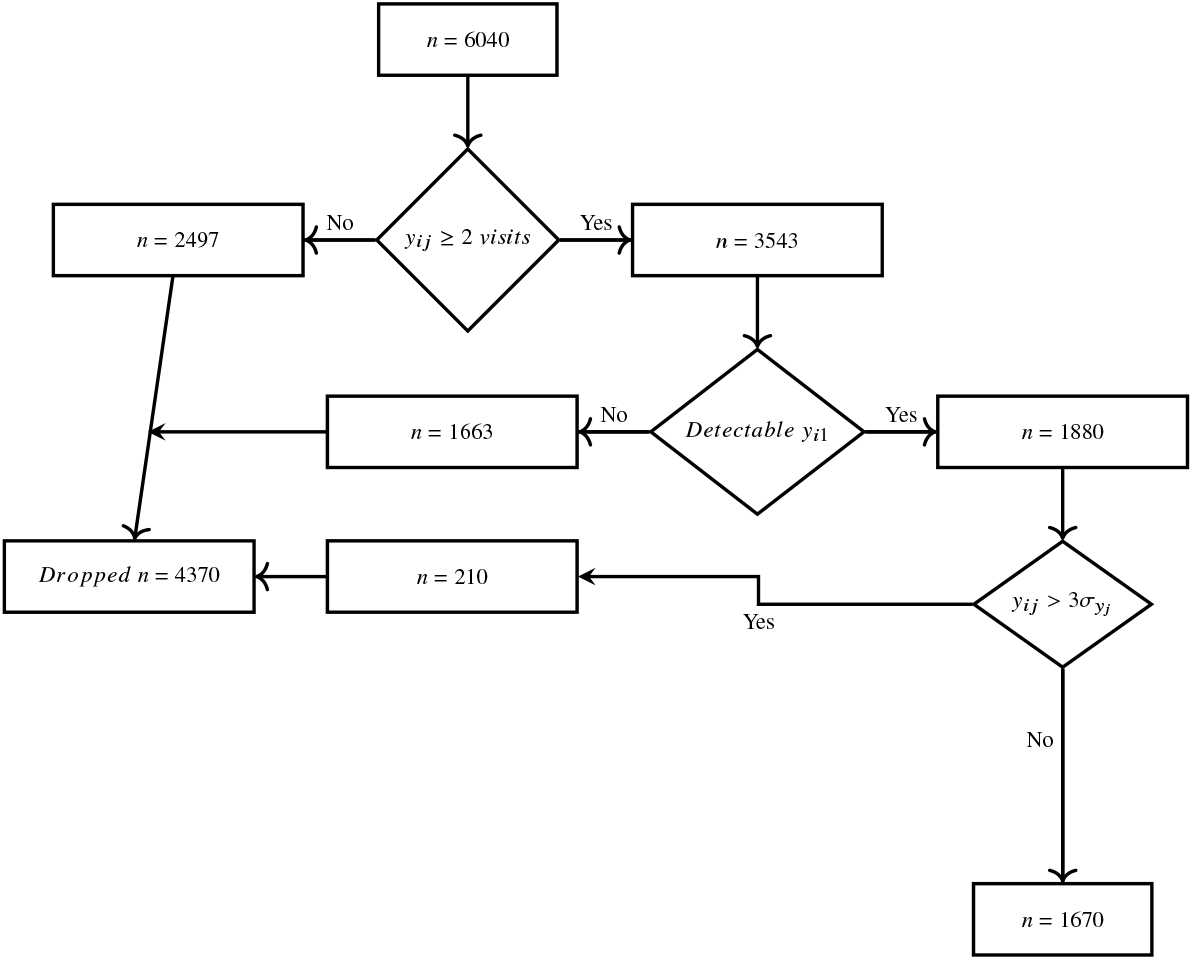
Inclussion and exclusion criteria

### Description of the variables

#### Dependent Variable

The outcome variable was the viral load copies measured as the number of ribonucleic acid per millitter of blood taken from HIV patients at a given visit. All viral load readings were transformed into one variable by reshaping the data and analyzed as a continuous outcome variable.

#### Independent variables

The independent variables extracted from facility databases are age, sex, marital status, treatment start date, patient’s last visit date, initial treatment type, current treatment type, adherence to treatment, patient’s baseline height and weight, and the World Health Organization’s (WHO) clinical stages.

From some of variables recieved, analysis variables were derived which include; age category, resulting from transforming numeric age into age groups guided by the baseline age of the patient; duration on treatment, obtained as the difference between treatment start date and last visit date; baseline Body Mass Index (BMI) obtained as the ratio of weight (in kilograms) to squared height (in square meters), *i*.*e*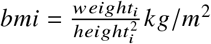; change in treatment regimen, obtained as logical test on whether a patient has ever changed treatment or not; and lastly, suppression history, which was also obtained as a logical test for each of the viral load measurements on whether the patient had less than 1000 copies/millitter of blood or not.

### Handling Missing Values

For this study, a total of 100 imputed data sets were generated by *mice*(), where each datum with missing information was imputed using the following methods: predictive mean matching (*PM M*) algorithm for all continuous variables; polytomous regression (*POLY REG*) was used for nominal data with more than two levels; logistic regression (*LOGREG*) was used for binary data; all of these methods were used within *M ICE* with twenty iterations [8]. No imputations were performed under SMF as all variables in this framework did not pass the inclusion criteria. Under RMF, a generalized linear mixed effect model was fitted to the gamma family and deployed on each of the 100 data sets with 30 iterations. At the pooling stage, *M ICE* selects the best model based on the smallest Bayesian Information Criterion (*BIC*), suggested by [1] as shown by **Equation 1**.

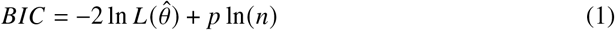

Where

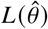 is the likelihood of the estimated model (covariates)

*p* is the total number of parameters that are estimated in the model

*n* is the sample size

#### Model Specification

Prior to model fitting, the data was reshaped into a long format, with repeating variables being viral load copies taken at each of the visits. After reshaping, a total of 10,014 records were generated, and a generalized linear mixed model (*GLM M*) was fitted with the generated viral load variable as the dependent variable and Socio-demographic, and Clinical factors as independent variables.

The models estimated in this framework were based on model separations defined by [14] and [3]. It was assumed that viral load copies vary by health facility, and hence the variable was used as the random effect term in the modeling process.

Let *Y*_*i j*_ be the *j* ^*th*^ VL copy for *j* = 1, 2, …, *n*_*i*_ from cluster (facility) *i i* = 1, 2, …, *m*. Suppose **X** is an *nm* × *p* fixed-effects design matrix. If *Y*_*i j*_ given **X** and *b*_*i*_ are independent random variables in a natural exponential family.

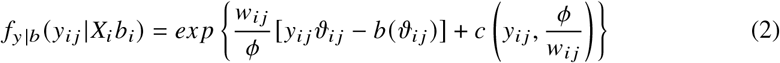

*Where*

*θ* and *ϕ* are unknown parameters to be estimated, *b and c* are known functions *w*_*i j*_ are known weights.

If

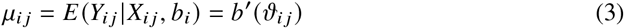

The generalised linear mixed model for *Y*_*i j*_ is give by

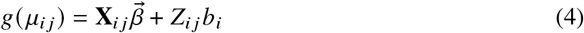

*g*(*μ*_*i j*_) is the link function, 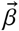 is a *p* × 1 vector of model parameters, **X**_*i j*_ fixed-effects design matrix, *Z*_*i j*_ is the random effect term (*health facility in this case*), *b*_*i*_ *∼ f* (*b*_*i*_ |*ψ*), and *f* (.|*ψ*) is a regular joint density for the vector of random effects, and *ψ* is the variance component of the random effect.

It is assumed that

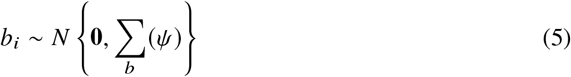

*Where*

**0** and Σ_*b*_ (*ψ*) are the mean vector and variance covariance matrix of the random effect respectively

### Ethical Considerations

Ethical review was saught from Makerere University’s College of Business and Management Science Research and Ethics Committee (CoBAMS-REC), review number CoBAMS-REC-2023-2. The study was retrospective in nature were neither study participants nor their spacemens were traced but rather their clinical characteristics recorded at the time of the viral load measurements/follow-up. Based on this, CoBAMS-REC waived the requirement of seeking consent from the study participants.

After ethical approval from CoBAMS-REC, a request for the data was made to the health facilities providing HIV treatment and care through the District Health Officer. Up-on approval by the medical directors of the study health facilities, data points including demographic, antiretroviral therapy (ART), clinical, and viral load information, were extracted from the Electronic Medical Records (EMR) databases of the facilities. Importantly, the data extracted from these systems was devoid of any personal patient information, ensuring the privacy of research participants during subsequent analysis. The study was conducted with complete transparency regarding its primary objectives and was free from any deceptive practices.

Furthermore, no affiliations, funding sources, or conflicts of interest that could potentially introduce bias into the study’s findings were declared.

### Limitations of the Study

The study relied on routinely generated data from healthcare facilities for its analysis, providing convenience but raising significant concerns about data quality. The absence of crucial variables, such as patient education level, in the datasets from these facilities posed a limitation. This limitation could hinder the study’s ability to comprehensively understand the contextual factors influencing viral load copies, highlighting the importance of acknowledging and addressing the limitations associated with data quality and completeness.

## Results

### Summary Statistics

From **Table 1** the youngest person included in the study was 3 years old, while the oldest was 82 years old, and the average age was 37 years. On average, Patients spent 46.79 months, with the longest duration of 87 months (7.25 years). The highest body mass index (BMI) was 1083.68*kg*/*m*^2^ with an average of 24.54*kg*/*m*^2^. Majority were female patients (67.01%), between 15 and 49 years (84.13%), currently married (53.86%), and accessing treatment from a private facility (57.37%). Most of the patients had never changed treatment (59.77%), and were at the first stage of the WHO clinical stages (80.54%). For covariate information, variables with missing values included marital status, duration of treatment, adherence to ART, body mass index, and WHO clinical stages. There were no missing cases for the first time of measurement, but all other visits included in the analysis had at least 15% missingness. Overall, missing data was 18.5% of the entire data set.

**Table 1.** Summary of continuous variables.

| Summary Statistic | Age | Duration on Treatment | Body Mass Index |
| --- | --- | --- | --- |
| Minimum | 3 | 14 | 0 |
| Lower Quartile | 29.25 | 26 | 18.86 |
| Median | 35 | 47 | 22.22 |
| Mean | 36.78 | 46.79 | 24.54 |
| Upper Quartile | 43 | 67 | 26.14 |
| Maximum | 82 | 87 | 1083.68 |

**Source:** Generated from research data

**Table 2** presents the distribution of viral load copies at each time of measurement. The highest viral load copies (4793 *copies*/*ml of blood*) were taken at the second visit. All patients managed to have their copies assessed at the first time of measurement, and almost all (99.22%) had achieved a suppression status. At all other times of measurement, the majority of the patients that managed to have their copies assessed were able to receive a suppression status.

**Table 2.** Summary of numerical viral load copies.

| Summary Statistic | Viral1 | Viral2 | Viral3 | Viral4 | Viral5 | Viral6 |
| --- | --- | --- | --- | --- | --- | --- |
| Minimum | 0 | 0 | 0 | 0 | 0 | 0 |
| Lower Quartile | 50 | 30 | 0 | 29 | 0 | 0 |
| Median | 50 | 68 | 0 | 29 | 50 | 0 |
| Upper Quartile | 55 | 70 | 0 | 57 | 50 | 57 |
| Maximum | 2400 | 4793 | 1940 | 76 | 289 | 2747 |

**Source:** generated from research data

### Inferential Results

At this level of analysis, the aim was to assess the effect of patient covariates on viral load copies using only variables with a *p* < 0.2 at the exploratory stage. The analysis was performed only for the repeated measures framework since no variable had a significant effect on viral load copies under the single measures framework. For the RMF, all variables had a significant effect on viral load copies except body mass index, and hence all were included in the modeling process. This section presents the results of a generalized linear mixed model with a gamma family before and after imputations are reported and discussed.

Panel A of Table **3** present the results of the generalized linear mixed models (*GLM M*) fitted on repeated measurements of viral load copies, while Panel B presents the pooled results of the same model fitted on the 100 imputed data sets resulting from multiple imputation (MI) with 30 iterations. The model fitted to multiply imputed data generated results slightly different from that fitted under complete case analysis. In both models, patients’ marital status, the facility from which a patient access treatment, and the duration on treatment had a significant effect on viral load copies. Patient’s gender, age, change in treatment, adherence rating, and WHO clinical stages had no effect on viral load copies.

**Table 3.**
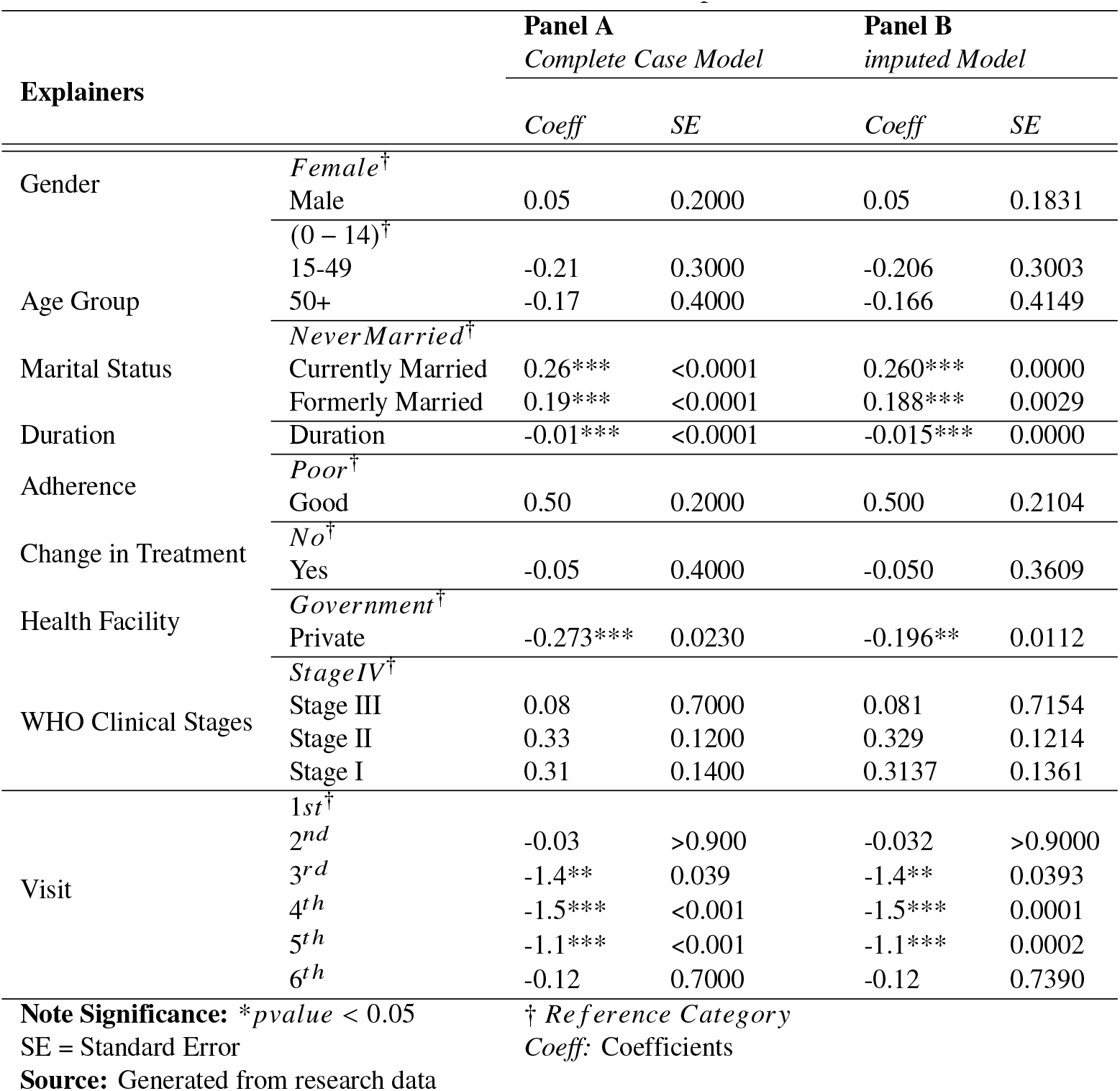
Model estimation under RMF before and after Imputaion.

| Explainers |  | Panel A<br>*Complete Case Model* |  | Panel B<br>*imputed Model* |  |
| --- | --- | --- | --- | --- | --- |
|  |  | *Coeff* | *SE* | *Coeff* | *SE* |
| Gender | *Female*<sup>†</sup> |  |  |  |  |
|  | Male | 0.05 | 0.2000 | 0.05 | 0.1831 |
| Age Group | (0 – 14)<sup>†</sup> |  |  |  |  |
|  | 15-49 | -0.21 | 0.3000 | -0.206 | 0.3003 |
|  | 50+ | -0.17 | 0.4000 | -0.166 | 0.4149 |
| Marital Status | *NeverMarried*<sup>†</sup> |  |  |  |  |
|  | Currently Married | 0.26*** | <0.0001 | 0.260*** | 0.0000 |
|  | Formerly Married | 0.19*** | <0.0001 | 0.188*** | 0.0029 |
| Duration | Duration | -0.01*** | <0.0001 | -0.015*** | 0.0000 |
| Adherence | *Poor*<sup>†</sup> |  |  |  |  |
|  | Good | 0.50 | 0.2000 | 0.500 | 0.2104 |
| Change in Treatment | *No*<sup>†</sup> |  |  |  |  |
|  | Yes | -0.05 | 0.4000 | -0.050 | 0.3609 |
| Health Facility | *Government*<sup>†</sup> |  |  |  |  |
|  | Private | -0.273*** | 0.0230 | -0.196** | 0.0112 |
| WHO Clinical Stages | *StageIV*<sup>†</sup> |  |  |  |  |
|  | Stage III | 0.08 | 0.7000 | 0.081 | 0.7154 |
|  | Stage II | 0.33 | 0.1200 | 0.329 | 0.1214 |
|  | Stage I | 0.31 | 0.1400 | 0.3137 | 0.1361 |
| Visit | *1st*<sup>†</sup> |  |  |  |  |
|  | 2<sup>nd</sup> | -0.03 | >0.900 | -0.032 | >0.9000 |
|  | 3<sup>rd</sup> | -1.4** | 0.039 | -1.4** | 0.0393 |
|  | 4<sup>th</sup> | -1.5*** | <0.001 | -1.5*** | 0.0001 |
|  | 5<sup>th</sup> | -1.1*** | <0.001 | -1.1*** | 0.0002 |
|  | 6<sup>th</sup> | -0.12 | 0.7000 | -0.12 | 0.7390 |

**Note Significance:** \**pvalue* < 0.05
SE = Standard Error
**Source:** Generated from research data<sup>†</sup> *Reference Category**Coeff*: Coefficients

From the table, for both methods of dealing with missing values, currently or formerly married patients were likely to have higher viral load copies (*p* < 0.05). For clinical factors, duration on treatment, and attending treatment from a private health facility were both associated with lower viral load copies (*p* < 0.05) for both methods of dealing with missingness. The model based on imputed data indicates a further reduction in the viral load copies for patients attending treatment at a private facility compared to the complete case model.

The table also indicate a significant reduction in viral load copies over time from the third vist to the fifth visit compared to the first visit (*pvalue* < 0.05) . The second and the sixth visits had no significant differences in viral load copies compared to the first visist (*pvalue* > 0.05)

## Discussion of Results

Panel B of Table **3** presents the aggregated outcomes derived from the Generalized Linear Mixed Model (*GLMM*) fitted to the 100 imputed datasets generated through multiple imputation (MI). From the model, no significant effect was established on patient gender and viral load copies(*pvalue* > 0.05), despite the variable manifesting individual significance in the exploratory model. This reinforces the need for access to treament and care with out gender baises in order to reduce the transmission rates of HIV. Furthermore, adherence to Antiretroviral Therapy (ART) did not emerge as a significant predictor of viral load (*p* > 0.05), consistent with results repoted by [4] when exploring adherence as a mediating factor between psychosocial variables and HIV viral load copies. It is also worth noting that literature has strong support for good adherence rating for HIV treatment to achieve the desired goal of zero transition rate.

### Distribution of the Random effect Term

Exploratory analysis established evidence variation in viral load copies at health facilitly level, as a result, the factor (health facility) was adopted as the random effect variable in the analysis. The results below are the random effect terms from all the 100 imputed data sets.

**Table 4** shows the variations in viral load copies across health facilities over time. The first row indicates an overall variation in viral loads of 2.34414, suggesting variability in the copies of viral loads across health facilities. Subsequent rows present for the random effects associated with each visit. Where *corr* = −1 indicates there is a perfect negative correlation in viral loads between that visit and the previous vist and the reverse is true where *corr* = 1. For example *corr* = 1 for the 3^*r d*^ visit indicates that on the third visit, vrial loads copies are increasing in HIV patients irresepective of where they are accessing treatment. A similar interpretation can be infered in cases where *corr* = −1. Overall, the table offers insights into the variability of viral load copies across health facilities and the correlations between viral load copies for different visits.

**Table 4.**
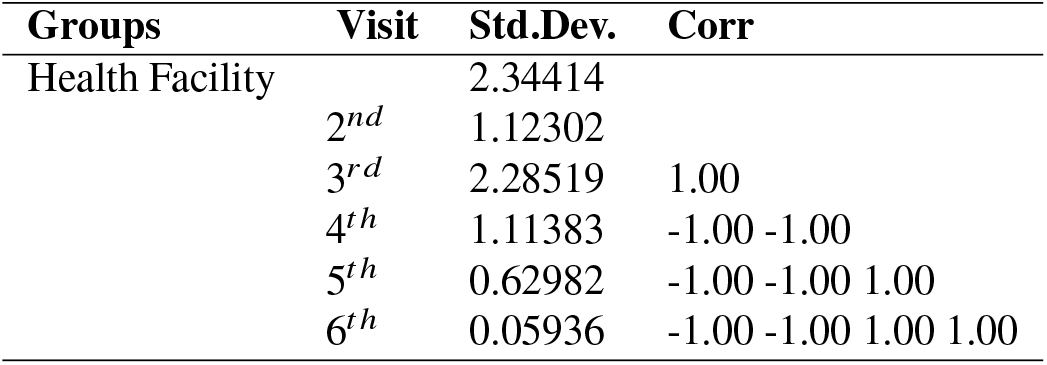
Random Effects.

| Groups | Visit | Std.Dev. | Corr |
| --- | --- | --- | --- |
| Health Facility |  | 2.34414 |  |
|  | 2<sup>nd</sup> | 1.12302 |  |
|  | 3<sup>rd</sup> | 2.28519 | 1.00 |
|  | 4<sup>th</sup> | 1.11383 | -1.00 -1.00 |
|  | 5<sup>th</sup> | 0.62982 | -1.00 -1.00 1.00 |
|  | 6<sup>th</sup> | 0.05936 | -1.00 -1.00 1.00 1.00 |

## Conclusion

The best-selected model was a generalized linear mixed model fitted to multiply imputed data. This aligns well with the study’s objectives and hypotheses, suggesting that a repeated measures framework and multiple imputation outperform a single measures framework and complete case analysis in assessing patient factors affecting viral load copies in longitudinal settings. The study also concluded that patient gender and adherence rating have no significant effect on viral load copies. Additionally, a significant reduction in viral load copies over time was observed in this study.

## Recommendations

The results of the study indicate the importance of using all information collected in the repeated measures setting and stresses the importance of carefully handling missing values in repeated measures data rather than dropping all records with missing observations from the analysis. It also advocates for the adoption of repeated measures analysis in HIV research to establish the magnitude of viral load copies and associated factors in people living with HIV.

Because the data utilized in this study was gathered as routine activities at a health facility, it’s possible the quality of the data may have affected the outcomes of the applied models. It is relevant to assess the repeated measures framework and the single measures framework on data collected under controlled conditions. Additionally, the study suggests implementing intervention programs for patients with marital histories (currently married and formerly married) to ensure optimal treatment response. Government health facilities should enhance HIV care services, potentially achieved through oversight and offering incentives to ART staff.

## Acknowledgments

We are grateful to the medical directors for their approval to use data at their health facilities and the invaluable efforts of the records departments during the data extraction process. Their contributions were essential to the success of this study.

## Data Availability

The data used in this study can be requested from the health facilities providing HIV treatment and care in Mukono district, specifying the period between January 2016 and December 2022. The District and/or health facilities reserve the right to grant access to the data.

